# The global scientific research response to the public health emergency of Zika virus infection

**DOI:** 10.1101/19001313

**Authors:** Juliane Fonseca de Oliveira, Julia Moreira Pescarini, Moreno de Souza Rodrigues, Bethania de Araujo Almeida, Claudio Maierovitch Pessanha Henriques, Fabio Castro Gouveia, Elaine Teixeira Rabello, Gustavo Correa Matta, Mauricio L. Barreto, Ricardo Barros Sampaio

## Abstract

**Background:** Science studies have been a field of research for different knowledge areas and they have been successfully used to analyse the construction of scientific knowledge, practice and dissemination. In this study, we aimed to verify how the Zika epidemic has moulded scientific production worldwide analysing international collaboration and the knowledge landscape through time, research topics and country involvement.

**Methodology:** We searched the Web of Science (WoS) for studies published up to 31st December 2018 on Zika using the search terms “zika”, “zkv” or “zikv”. We analysed the scientific production regarding which countries have published the most, on which topics, as well as country level collaboration. We performed a scientometric analysis of research on Zika focusing on knowledge mapping and the scientific research path over time and space.

**Findings:** We found two well defined research areas divided into three subtopics accounting for six clusters. With regard to country analysis, the USA followed by Brazil were the leading countries in publications on Zika. China entered as a new player focusing on specific research areas. When we took into consideration the epidemics and reported cases, Brazil and France were the leading research countries on related topics. As for international collaboration, the USA followed by England and France stand out as the main hubs. The research areas most published included public health related topics from 2015 until the very beginning of 2016, followed by an increase in topics related to the clinical aspects of the disease in 2016 and the beginnings of laboratorial research in 2017/2018.

**Conclusions:** Mapping the response to Zika, a public health emergency, demonstrated a clear pattern of the participation of countries in the scientific advances. The pattern of knowledge production found in this study represented the different perspectives and interests of countries based firstly on their level of exposure to the epidemic and secondly on their financial positions with regard to science.

## Introduction

Zika virus infection (ZVI) was first reported in humans in 1954, but only in 2007 did the first outbreak occur in Micronesia [1, 2], and several small outbreaks in Micronesia and the Pacific Islands have been reported since then. However, it was only in 2013 that ZVI started revealing its epidemic potential [3]. In 2015, the virus was linked with associated neurological disorders, including Guillain-Barré syndrome, microcephaly and yet unknown neurological disorders in newborns [4, 5]. It was and remains an event of scientific uncertainties.

In February 2016, mostly driven by the Brazilian Zika-associated microcephaly cases, WHO declared a public health emergency of international concern (PHEIC). Different research groups all over the world, as part of separate and collaborative research networks, made a concerted effort to understand the various factors required to cope with the epidemic, including the Zika virus infection, transmission and control, as well as the social impact of severe forms of the disease in the most affected societies [3]. During the outbreak, especially after WHO PHEIC, international funding and fast-tracking procedures for publishing scientific outcomes stimulated scientific research and publication. The increasing number of publications on Zika after 2015 has been attributed mainly to its association with microcephaly [6], but by 2016, only a few core journals had published on Zika [7].

Since the emergency declaration, science studies’ applied to the Zika epidemics publications has come mostly from focusing on scientific networks and their collaboration and co-publication. Some researchers have adopted bibliometric and/or scientometric perspectives, analysing papers and documents to discuss different aspects of scientific production in terms of co-authorship, cooperative networks and innovative potentials on the Zika topic (see [7-10]).

Furthermore, the discussion about the influence and centrality of certain groups, researchers and even countries have enriched the debate raised by scientometrics applied to Zika. A first mapping of the scientific networks and their most influential researchers in Zika as a theme was done in 2018 [11]. Using techniques of Social Network Analysis, the authors mapped the co-authorship networks on Zika papers published in 2015 and 2016. The study revealed that, besides the number of publications, two other factors might be connected to explain the prominence of a given researcher in these scientific networks: the diversity of partnerships and the established connections with the pioneers in the field.

Another interesting perspective is to address the role of specific regions on the matter. Zika-related publications and patents, investigated from a scientometric perspective, highlighted the strategic role played by the Latin America and Caribbean research network in terms of addressing the science and technology (S&T) challenges related to the outbreak [12]. According to this study, Brazil itself stands out in reporting and researching the clinical manifestations of the ZVI and also in studies about vector control, the latter related to the filing of patents in particular.

State of the art considered, these studies are central to the systematisation of current knowledge in terms of which research areas have been most developed so far. This scenario allows, for instance, policy managers to contribute by allocating and prioritising research to the current prevention and epidemiological combat of the Zika virus. It is also essential to identify interaction and collaboration among countries to help further the development of shared governance in Zika and other research-related fields and increase scientific production and knowledge in the area. Regarding neglected populations and diseases, these studies can support further analysis on equity and sustainability, and science production between low and middle-income countries and developed countries, during and after health emergencies.

In this study, we aimed to verify how the Zika epidemic has moulded scientific production worldwide analysing international collaboration and the knowledge landscape through time, research topics and country involvement. Therefore, this study explores how the epidemic of Zika has translated into research all over the world and over time, identifying what the main thematic areas of scientific development on Zika are. We also explore how the S&T apparatus of each country responded to the Zika emergency, from the first studies until 2018 and according to knowledge areas.

## Materials and methods

### Data collection, treatment and cleaning

Our dataset was obtained by searching terms related to Zika virus infection (“zika” OR “zkv” OR “zikv”) by Topic in Web of Science (WoS). We selected all the articles published from 1945 to the end of December 2018. The search was performed on January 21st, 2019. From the retrieved articles, we extracted the title and abstract to develop a knowledge map and used the metadata variables regarding date published, authors and co-authors’ affiliation countries as well as the ISI subject categories from WoS assigned to them [13].

The selected contents were downloaded as a text file containing all the results of our query. The file was cleaned by (i) converting all words to lowercase, (ii) removing special characters (e.g., “.”, “:” and “/”), and (iii) standardizing similar terms (e.g., “ae aegypti”, “aedes aegypti”). Data cleaning was conducted in Python. The Python code used is available at GitHub [14].

### Data Analysis

We conducted two main analyses on the Zika publications retrieved. First, a descriptive analysis to understand how countries collaborate towards Zika knowledge; and second, a textual analysis of research on Zika using knowledge mapping over time and space (i.e., countries).

#### Descriptive analysis

The descriptive analysis was performed both directly by filtering information from Web of Science homepage and by using VOSviewer software [15] to perform country collaboration network analysis. To summarize data information for each research article, we extracted from each author his institutional affiliation country. Articles produced by different institutions of the same country were considered a national collaboration, and articles produced by authors affiliated to institutions from more than one country were considered international collaborations. After that, we constructed a co-authorship network at the country level using data exported from VOSviewer and reconstructed and filtered using Gephi software for visualization [16].

In order to measure collaboration in Zika publications, we deduced an international collaboration index (ICI), which is the ratio of the number of countries that a given country collaborated with, over the total number of countries (minus the country itself) which produced results toward Zika knowledge. The closer the index is to one, the more collaborative the country is, and the closer it is to zero, the less internationally collaborative the country is.

To address the issue of international collaboration over time as a proportion of published articles, an international collaboration factor (ICF) was calculated as the number of articles with author(s) affiliation from more than one country over the number of articles with author(s) affiliation to just one country. As with the ICI, the more colaborative a country is, the closer it is to one, the less collaborative, the closer it is to zero.

#### Textual analysis

We carried out a textual analysis to discover textual data relationships among extracted words. The method reveals thematic trends and identifies publication subjects surrounding specific topics or fields [17]. To do this, we analysed the titles and abstracts of the articles in our sample. More specifically, we analysed words (also called terms or forms), extracted from the titles and abstracts, grouped by text segments comprised of 40 terms each, and applied Correspondence Factorial Analysis (CFA) to correlate them. The baseline assessment of research areas into clusters relied on the clustering algorithm of IRAMuTeQ v7.2 (Interface for Multidimensional Analysis of Texts and Questionnaires). The correlation process is obtained by testing if the frequency of a given word is statistically associated with another using Pearson Chi-square at a significance level of 5%. Based on the similarity, words are aggregated into clusters of main areas of knowledge. Content analysis was also performed using IRAMuTeQ.

The defined number of clusters in IRAMuTeQ required empirical knowledge in the field as well as several tests on how many clusters made a fair and accurate separation of knowledge areas. Therefore, to validate our baseline clusters, we compared the number and content of each cluster obtained using IRAMuTeQ with those obtained using VOSviewer. Moreover, we used the categories defined by WoS for each article, based on the journal in which it was published to further evaluate if the clusterisation process and defined sub-areas of research were aligned. Finally, we showed the results to a panel of specialists on Zika and public health.

After we had established statistically significant knowledge area clusters, we evaluated which knowledge areas emerged over time, and stratified them into four groups by publication year: <=2015, 2016, 2017 or 2018; second, we evaluated the participation in each cluster/field of the top 10 countries with regard to the number of publications.

## Results

### Descriptive analysis on the countries collaborations

The collected data yielded a total of 3,454 articles produced by 18,453 authors, associated with institutions from 150 countries. These articles were published with and without international collaboration. Fig 1 shows a dispersion matrix using the number of publications and ICI by country.

**Fig 1.**
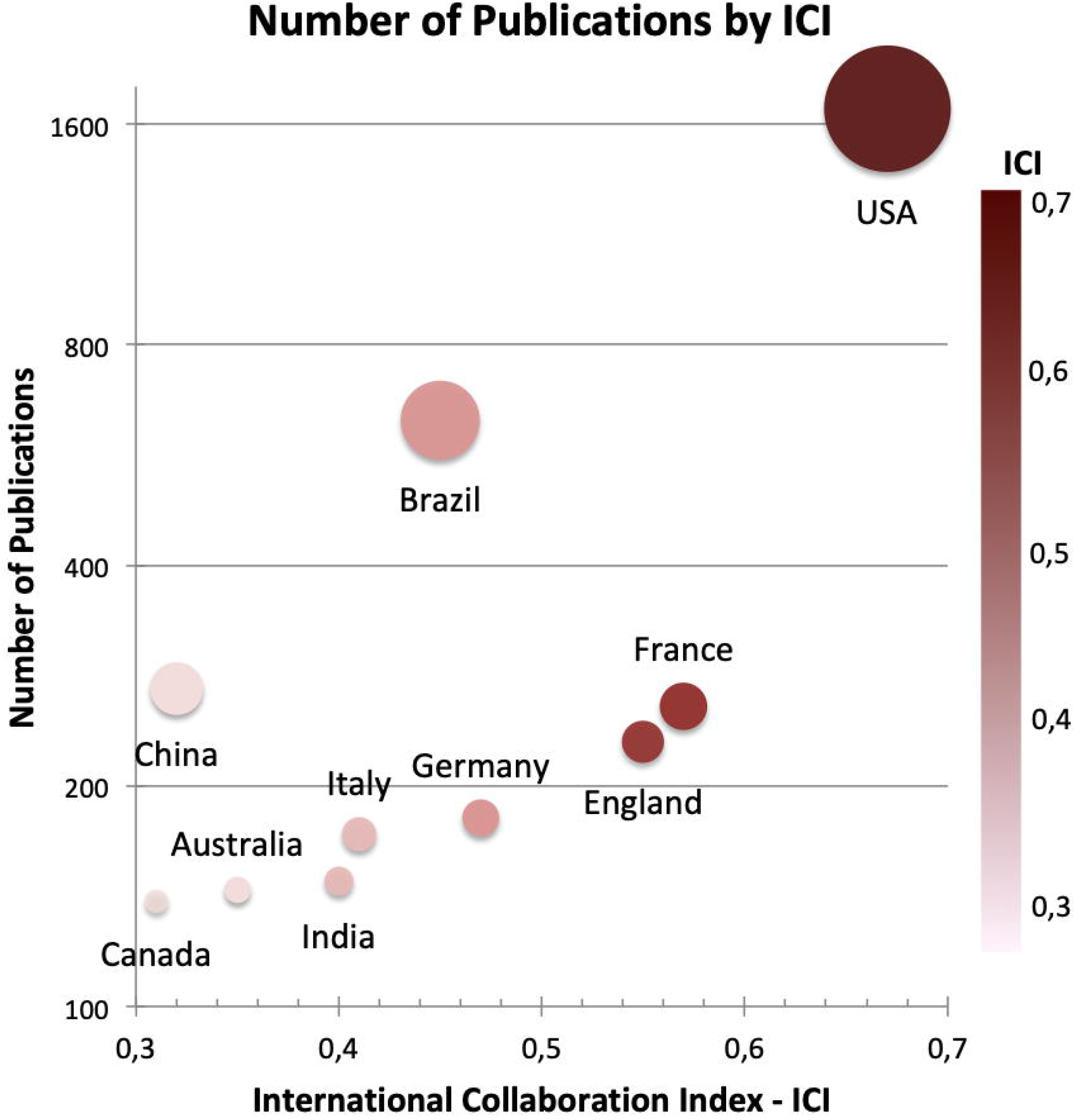
Number of publications by International Collaboration Index for each of the top ten countries. The graph is a dispersion matrix, the y axis is the number of publications on a logarithmic scale base 2 starting at 100 publications and the ICI is on the x axis starting at 0.3. The size of the circles is related to the number of publications and the intensity of the color from dark red to light pink is related to the ICI as shown on the right side scale.

According to Fig 1 The most internationally collaborative countries were the USA (ICI = 0.67), followed by France (ICI = 0.57), England (ICI = 0.55), Germany (ICI = 0.47) and Brazil (ICI = 0.45). The remaining most productive countries, Italy, India, Australia, China and Canada, presented international collaboration indexes of 0.41, 0.40, 0.35, 0.32 and 0.31, respectively. When we take into consideration the ICI with the number of publications we can see that the USA has the highest ICI and number of publications placing it in the top right corner of the graph. In contrast, China, the third country with the highest number of publications, is the ninth in ICI in the bottom left corner of the graph. Brazil seems to occupy an intermediate position regarding the number of collaborating countries and the number of publications, with the second largest number of publications on Zika, and the fourth in ICI. France and England seems to be well positioned in terms of the number of collaborating countries. The remaining five countries have fewer than 200 publications and show a similar ratio between the logarithmic number of publications and the number of countries with which they have collaborated.

Table 1 summarizes the overall information about international collaborations for the ten most productive countries. The countries that published the most on Zika during the study period were the USA and Brazil followed by China, France and England.

**Table 1.**
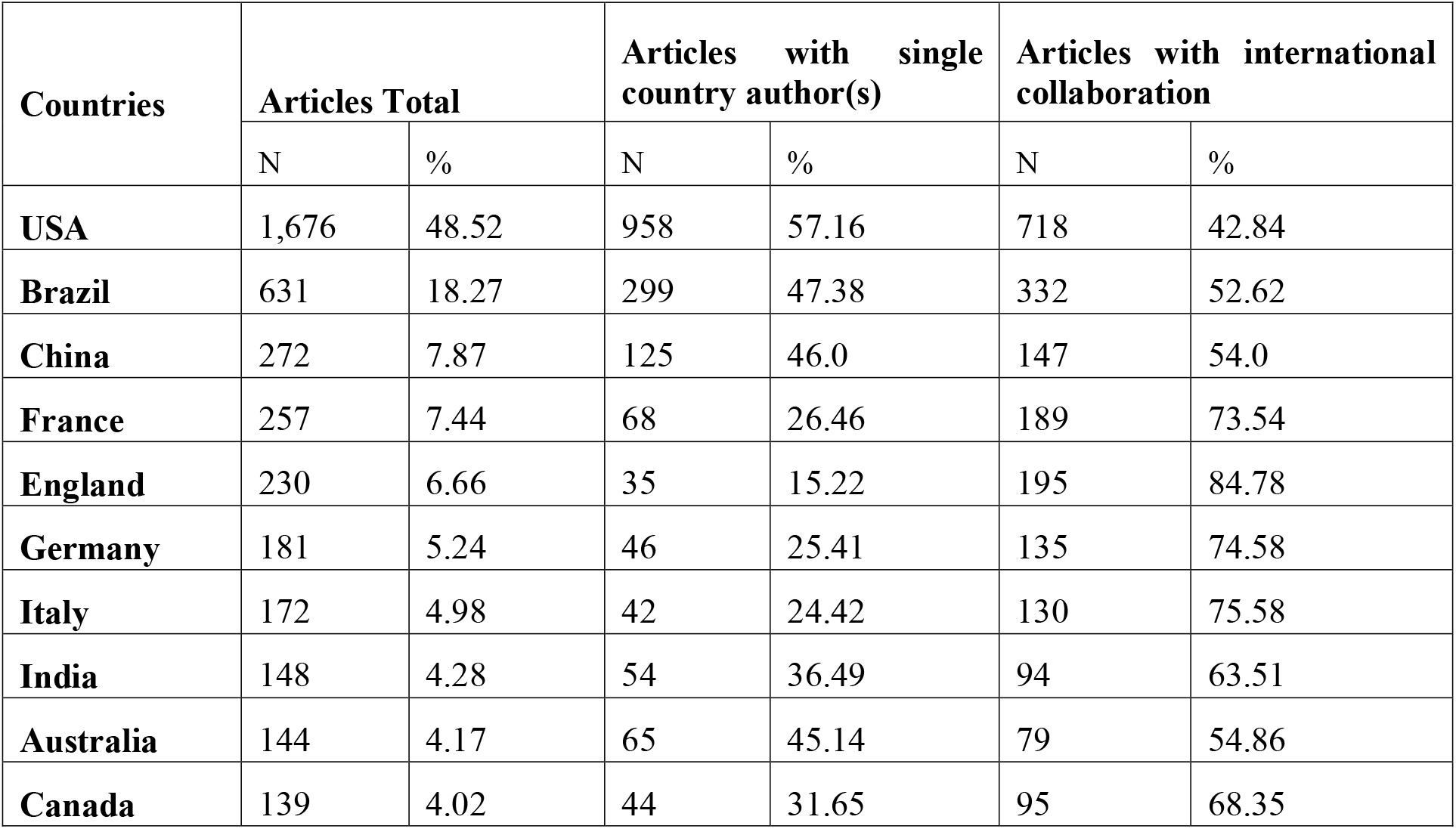
National and International publications about Zika Virus Infection for the top 10 most productive countries. The table depicts the total number of publications. The column “Articles Total” represents the total number of articles published by a country and the percentage of these productions over the total number of articles about Zika found in WoS until 2018. Among these values, we see the number of articles produced by the country itself and the proportion produced with international collaboration.

| Countries | Articles Total |  | Articles with single country author(s) |  | Articles with international collaboration |  |
| --- | --- | --- | --- | --- | --- | --- |
|  | N | % | N | % | N | % |
| **USA** | 1,676 | 48.52 | 958 | 57.16 | 718 | 42.84 |
| **Brazil** | 631 | 18.27 | 299 | 47.38 | 332 | 52.62 |
| **China** | 272 | 7.87 | 125 | 46.0 | 147 | 54.0 |
| **France** | 257 | 7.44 | 68 | 26.46 | 189 | 73.54 |
| **England** | 230 | 6.66 | 35 | 15.22 | 195 | 84.78 |
| **Germany** | 181 | 5.24 | 46 | 25.41 | 135 | 74.58 |
| **Italy** | 172 | 4.98 | 42 | 24.42 | 130 | 75.58 |
| **India** | 148 | 4.28 | 54 | 36.49 | 94 | 63.51 |
| **Australia** | 144 | 4.17 | 65 | 45.14 | 79 | 54.86 |
| **Canada** | 139 | 4.02 | 44 | 31.65 | 95 | 68.35 |

The column with the total number of articles and the percentage in Table 1 accounts for more than the total number of articles on Zika due to the collaboration amongst these countries. With regard to the articles written by authors from one country alone and with collaboration we can see that the only country with fewer than half of its articles being the result of collaboration is the USA, in contrast with England, which has published almost double that of the USA, with 84.78%. The other three European countries, France, Germany and Italy, also shows higher percentages compared to other non European countries.

In order to see how international collaboration among the top 10 publishing countries occurred over time, in Fig 2 we plotted the yearly evolution of their international collaboration factor - ICF (the number of articles produced with international collaborations over the total production) until 2018 combining papers before 2015 all together. We can see a clear pattern in the USA, Brazil, China and Australia, where the countries have a balanced level of production with and without international collaboration. As a contrast, we can see that England, Germany and France published articles in international collaborations at a higher frequency. Nonetheless, the yearly ICF of the ten most productive countries together with the ICF for Zika publication and WoS overall publication are considered in Fig 2.

**Fig 2.**
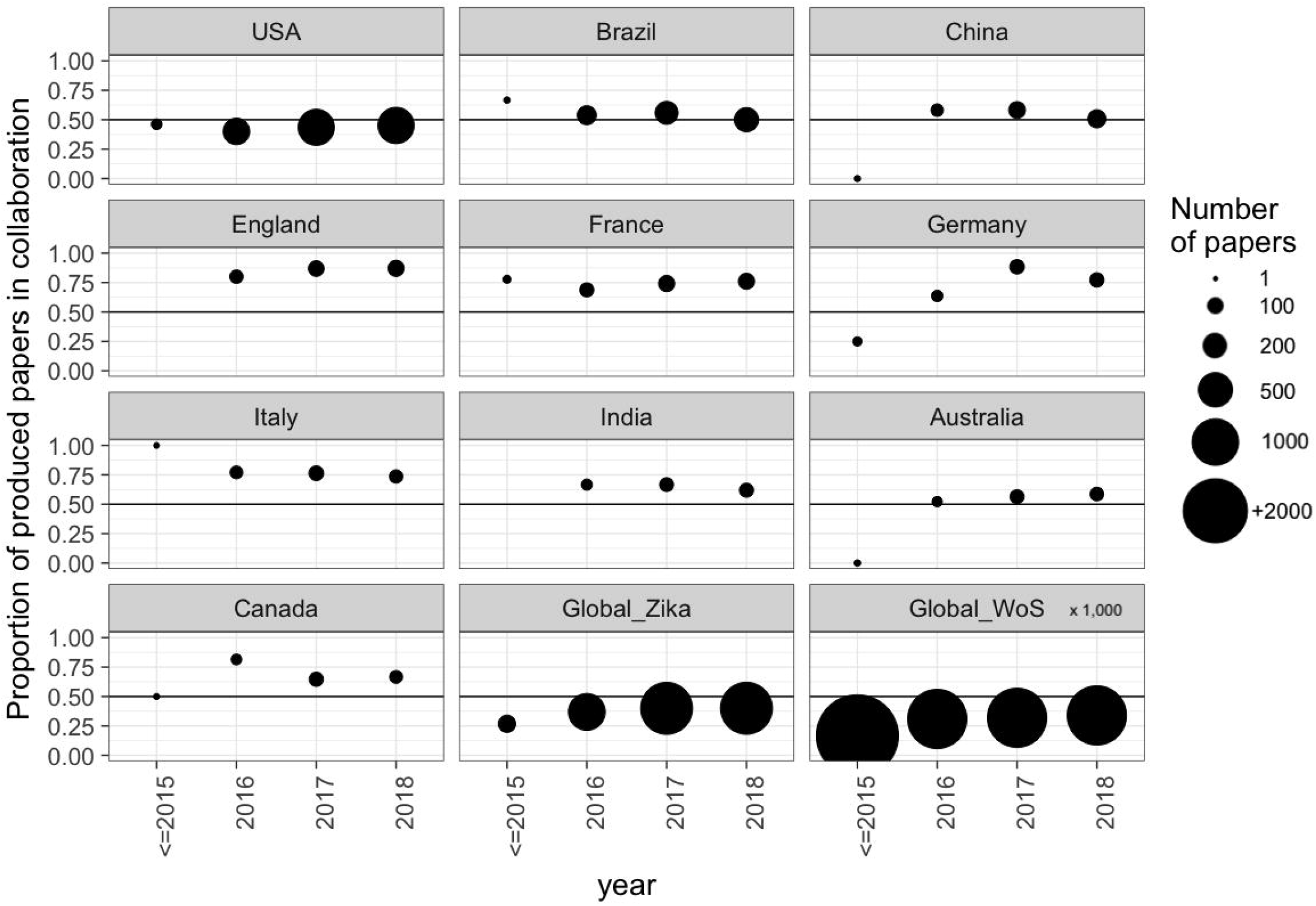
The production evolution of the top 10 countries plus the overall Zika Production and WoS production. The closer the dots are to 0.5 (highlighted by the black line), the articles produced with collaborations are proportional to the articles produced without collaborations. The value corresponding to the year 2015 represents the sum of all work produced by the selected country up until 2015. Points on the zero line correspond to publications that had no international collaboration. The size of the dots for Global WoS articles are divided by 1,000 in order to fit the scale.

Using co-authorship networks between countries, we see that the majority (61.89%) of the articles were produced within one country, 24.17% were collaborations between two countries, 8.88% between three countries and 2.51% between four countries. The remaining 2.55% were collaborations between five or more countries. Interestingly, we also found that international collaborations were present on average in 64.47% of articles produced by the top 10 countries. The reason for this apparent discrepancy has to do with the fact that an international paper will be considered a collaboration for every country represented in that paper but only once when comparing articles with and without international collaboration.

Analysing country-level collaboration, as shown in Table 1 England has the highest number of publications that resulted from international collaboration (84.78%), followed by Italy (75.58%) and Germany (74.58%). Another collaboration analysis which supports the results for Table 1 and Fig 2 is presented in Fig 3 with an international collaboration network graph. This figure presents all the collaborations amongst the top ten countries.

**Fig 3:**
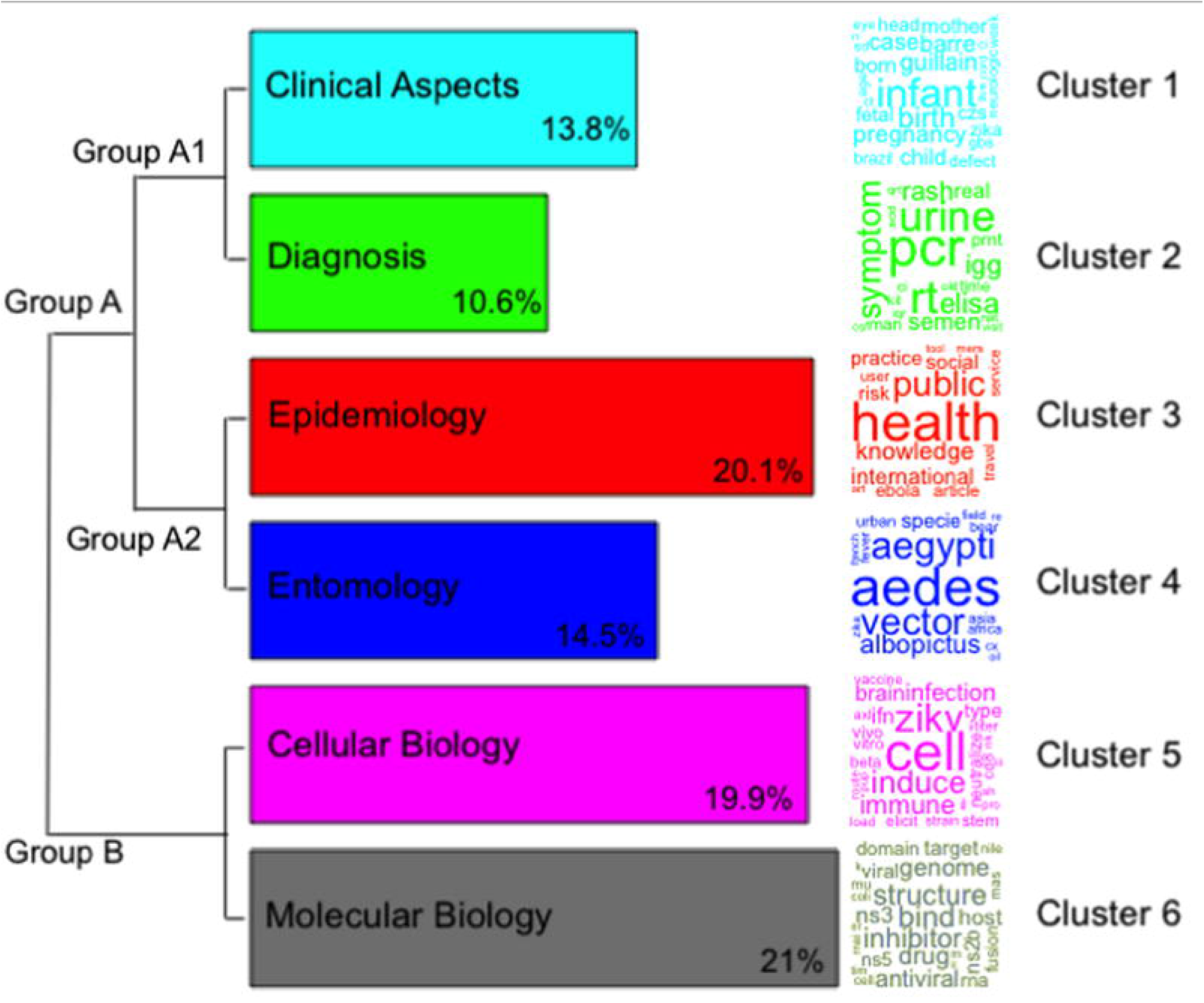
Collaborations among the 10 most productive countries. Each node corresponds to a country and the size of the node is related to the number of international collaborations they have within that group. The thickness of the lines represents the number of collaborations between any two countries and an aproximation of those numbers are presented by the box on the top right. The color scheme emphasizes link differences between countries since the color of a line is a mix of the color of the nodes it connects. The node distribution on the figure has been chosen only to facilitate visualization. The two countries with the fewest number of publications are positioned at the center of the graph and the most productive to the least from top to bottom and left to right.

The USA is in the first position in terms of absolute numbers of publications. In contrast, it occupies the 10th position (42.84%) in terms of the percentage of articles produced with international collaboration. Despite being in 10th position, the number of articles that the USA published with international collaboration corresponds to more than three times the total number of articles published by England, the country with the highest international collaboration ratio. Out of the 958 articles published by the USA with international collaboration, over 15% were with Brazil and nearly 10% with China. This strength of collaboration is shown in Fig 3 by the thickness of the line between the nodes.

Out of the 299 Brazilian studies published as results of international collaboration, over 60% were with the USA, followed by nearly 20% with England and 10% with both France and Germany. For the 147 Chinese studies published with international collaboration, over 75% were with the USA and a few articles with other countries. (Fig 3).

### Main research areas of knowledge

A total of 24,685 words were analyzed inside 21,055 text segments, of which 17,525 were considered active words (i.e., verbs, nouns and adjectives). Using IRAMuTeQ, six clusters were identified and named according to the most frequent words. Cluster 1 represented the “Clinical Aspects” of the disease (13.8% of the words); Cluster 2 the “Diagnosis” (10.6%); Cluster 3 included the “Epidemiology” of the disease (20.1%); Cluster 4 the “Entomology” (14.5%); Cluster 5 “Cellular Biology” (19.9%); and Cluster 6 “Molecular Biology” (21%) (Fig 4).

**Fig 4.**
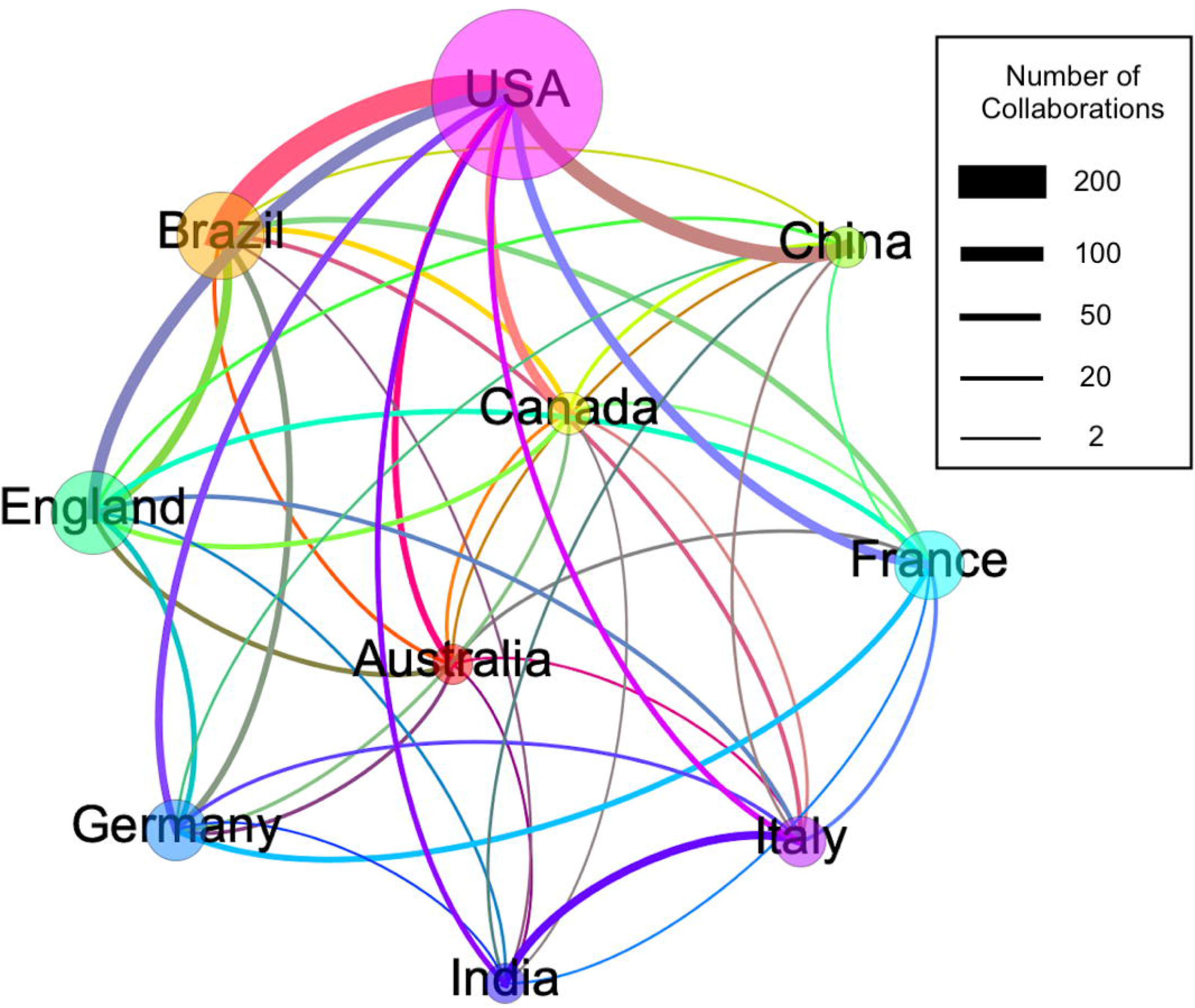
Dendrogram of the clusters obtained with IRAMuTeQ. The dendrogram is divided into 6 clusters formed by branch divisions. The first branch divides Group A, and Group B. Group A is further divided into two subgroups called Group A1 and Group A2. The size of each bar corresponds to the number of words in that cluster directly related to the percentage value in the bottom right corner of each bar. The associated name for each cluster appears on its bar, and the colour scheme has been chosen to facilitate visualization. In front of each bar, there is a square word cloud of words with the highest chi-square value to aid the understanding and the representation of each cluster.

Using the WoS subject categories, 165 of 252 possible categories were present in the studied articles, and each article is classified in one or more category. To compare WoS categories with the clusters defined using Iramuteq, we used the following approach. First, we selected the ten most frequent categories presented in our studied articles. Second, we analysed if the most significant categories for each one of the 6 clusters independently appeared with a high frequency or not. By doing this, we were able to better classify the clusters by the words which appeared in them.

The ten most frequent categories were: Infectious Diseases; Public Environmental Occupational Health; Tropical Medicine; Virology; Multidisciplinary Sciences; Microbiology; Parasitology; Immunology; Biochemistry Molecular Biology; and Medicine General Internal. Other six categories, among the 10% most frequent, which were not included in the top ten list but were significant to categorize the clusters were: Entomology; Cell Biology; Health Policy and Services; Clinical Neurology; Pediatrics; and Diagnosis. Based on these sixteen categories, we found the following results for Group A: Public Environmental Occupational Health category was well defined in the “Epidemiology” cluster (cluster 3). The clusters “Diagnosis” and “Entomology” (clusters 2 and 4) shared some of the same categories such as Infectious Diseases and Tropical Medicine, with a difference for the Parasitology category which appeared in cluster 4 but not in 2. Furthermore, Diagnosis category was in cluster 2 and Entomology category in cluster 4; each cluster was named after these categories. Cluster 1 “Clinical Aspects” had none of the top 10 high-frequency categories of WoS in it. However, it included less frequent categories such as Clinical Neurology and Pediatrics.

In Group B, with clusters “Cellular Biology” and “Molecular Biology” (cluster 5 and 6), there were similarities in Virology and Microbiology as main categories. The main difference was that in cluster 6, the most common category was Immunology, and in cluster 2 Virology with Multidisciplinary Sciences were some of the areas. The category Cell Biology was included in cluster 5 which was named after it. Similarly, the category Biochemistry Molecular Biology was very evident in cluster 6.

After the validation of the clusters by the categories and their representations, we asked a total of 12 specialists to evaluate the clustering results and the nomenclature. All responses were positive in considering the clusters a well-defined set of research sub-areas within the field of Zika taking into consideration the words available and the categories. Spatial distribution of the words coloured according to the cluster was presented to the specialists with the inferred nomenclature and positive responses were received.

### Year and Country distribution by cluster

Another scientometric analysis, after the construction of the knowledge map and the validation of the clusters, concerns the global research path of science over time, based on the year of publication of the articles. Because a large number of papers were published from 2016 onward we decided to group all publications before and including 2015 together. These accounted for 3.2%. The publications of our sample yielded four groups (<=2015, 2016, 2017 and 2018). We carried out this investigation in the same way as we did with the subject categories, by evaluating how statistically relevant a specific time group were in one or more of the six clusters.

We incorporated the findings into Fig 5 with the boxes and using tilde (∼) in front of the year to emphasise the fact that the results are an approximation. This means that a particular cluster might have publications or words in all year groups, although its prevalence is in a particular quadrant position or year.

**Fig 5.**
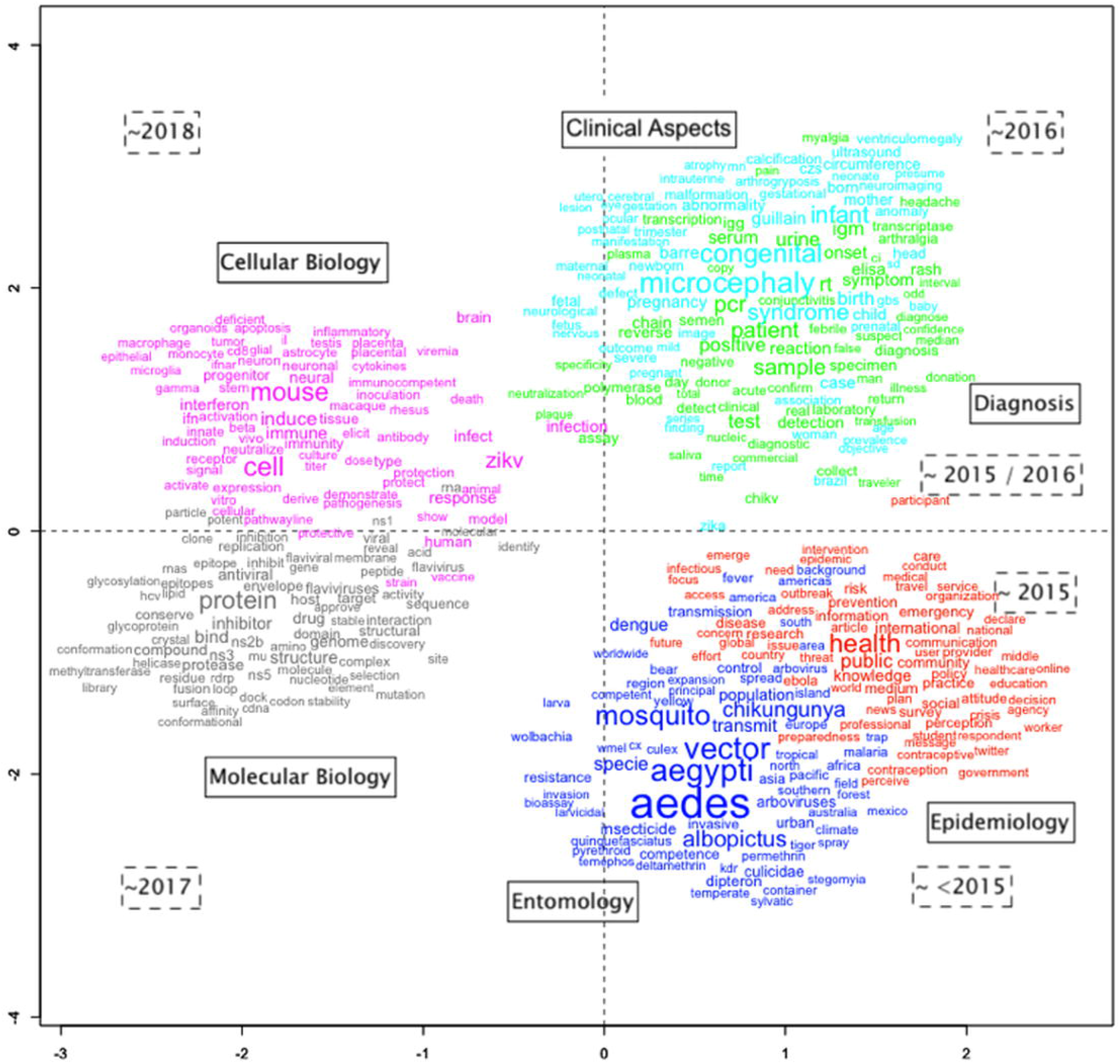
Knowledge map. Words extracted from titles and abstracts of articles on WoS related to Zika Virus infection from January of 1945 through December 2018. The names in the solid line boxes represent the cluster names and the dotted line boxes represent the most frequent periods for the articles represented by the clusters. The size of each word represents their chi-square value; the distance between them represents how statistically related they are based on the results of the correspondence factorial analysis.

Similar to the subject categories, the year of publication analysis presented a well-defined pattern. Publications before and up to 2015 were most prevalent in the bottom right part of the figure with entomology research and related words in years before 2015 and regarding public health in 2015 and 2016. Publications in 2015 showed a higher than expected presence, by analysing the chi-square, for clusters 3 “Epidemiology” and cluster 2 “Diagnosis”. For “Diagnosis”, the production surged between 2015 and 2016 the period at the start of the epidemics. The top right quadrant has publications from 2015 to 2016 mostly, where the clusters “Diagnosis” and “Clinical Aspects” are present. In broad terms, epidemiology is defined as the study of the distribution and determinants of health-related states or events in specified populations in order to characterise and determine associations between exposures and outcomes [18]. Taking into account that 2015 through 2016 was the most critical period due to knowledge gaps related to the infection, the three remaining areas in Group A - clinical aspects, diagnosis and entomology - are key factors to be studied in order to better understand epidemiology. Furthermore, entomology is related to the need for a public health quick response during an emergency.

In the left part of Fig 5, the clusters from Group B can be seen for both years 2017 on top and 2018 on the bottom. Each year is well established in those quadrants and aligned with the areas of research and clusters “Cellular Biology” and “Molecular Biology” respectively. Taking into consideration the areas with lower than expected values for chi-square for these years, cluster 4 “Entomology” is lower in 2017 and cluster 1 “Clinical Aspects” is lower for 2018, which shows a significant shift in research topics from previous areas to new areas.

Finally, we performed a country analysis for the clusters with the top ten countries regarding the number of publications. Fig 6 shows the Chi (not squared) results in order to capture the negative values (lower than expected) for the countries participating in a cluster. The higher or lower than expected results could be interpreted as the amount of research above or below average, represented on the text analysis, carried out by each country. If a country’s research distribution to a particular cluster is close to zero, the percentage of articles expected for that cluster follows the overall trend. If the number of articles is above or below the percentage expected for that cluster, a bar will appear considering how far from the expected value they are.

**Fig 6.**
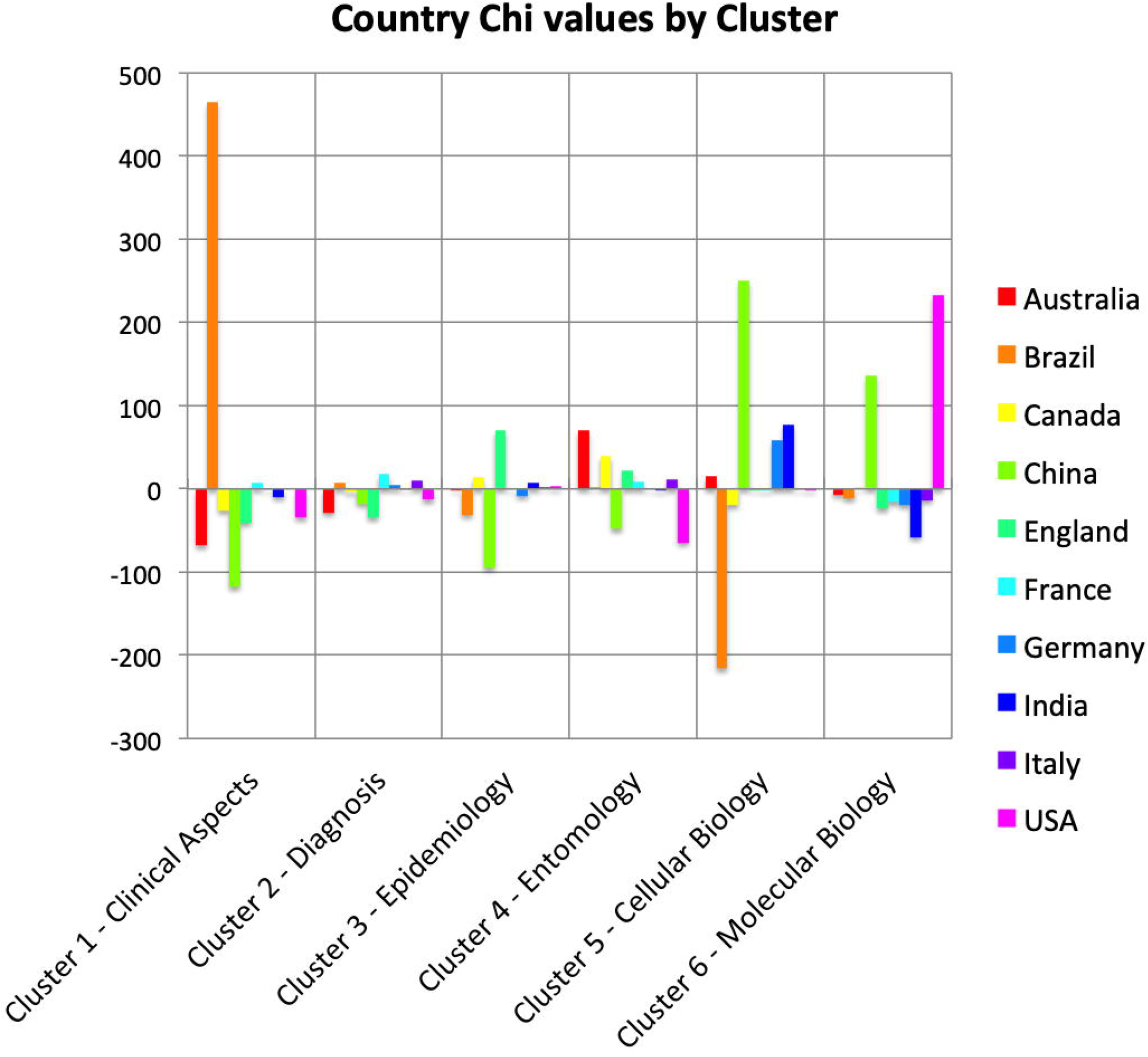
Chi values for the top 10 countries for each cluster. The results are calculated taking the observed value (number of text segments) within a cluster minus the expected value for that cluster and country divided by the square root of the expected value. This is the Chi value (not squared) to show negative (or less than expected) values. For each cluster (1 through 6) we have all ten countries and their respective chi value.

If a country shows a particular concentration in a cluster, it should show a lower than expected in one or more clusters. From Fig 6, we can infer the presence of a country as well as its contribution to research on the topic or area.

Starting with the most productive, the USA has a high presence with statistical relevance in cluster 6 focusing on “Cellular Biology”. Although the USA has a presence in all the 6 clusters, due to its high number of publications and collaborations, the most significant presence, statistically speaking, is in cluster 6 and its least significant presence in cluster 1 “Clinical Aspects” and 4 “Entomology”. Brazil, the country with the second highest scientific production is most significant in cluster 1 regarding “Clinical Aspects” of the disease, while it is almost absent in “Cellular Biology” (cluster 5). China is present in two clusters mainly, “Cellular Biology” (cluster 5) and “Molecular Biology” (cluster 6) along with the USA. Clusters 1 through 4 are of very little statistical relevance for China. France is present in clusters 1, 2 and 4 playing a role as an important research centre for public health but focusing on the disease as well since French Polynesia had been one of the first places to show endemic signs of the virus. About England, while a non endemic country, its important presence in cluster 3 “Epidemiology” might be related to the fact that it is home to one of the largest centre for tropical and neglected diseases in the northern hemisphere, the London School of Hygiene & Tropcial Medicine. Australia, Canada and Italy show a very high presence in cluster 4 “Entomology”. For Germany, India, and China, cellular biology (cluster 5) appears as the main area of research.

## Discussion

In this study, one of the first analyses on Zika publication that has taken into consideration international collaborations, we have attempted to understand the scientific research path towards Zika science production. The unexpected appearance of the virus in an epidemic form and with devastating effects on pregnancy was followed by the rapid growth of scientific research on ZVI with a great inflow of research capital and institutional interests. It makes research into Zika scientific production at the international context a compelling case in international health and biomedical research perspectives, given its unique characteristics.

In general, collaborations in science occur through the need for complementary resources among different actors, institutions and countries, such as knowledge and skills, financial resources and access to the field and key actors in order to conduct relevant research. Co-authorship studies indicate that more connected actors tend to attract a higher number of connections over time [19], an aspect that amplifies learning capacities and the absorption of knowledge. This study shows how ZVI related-science has evolved since 2015, as a result of the interest of an international research community focused on solving new and challenging research questions, related to a recognized worldwide health problem.

The appearance of ZVI in its epidemic form in the Americas in 2015, particularly in Brazil, initially triggered epidemiological field studies. In 2016, when the outbreak was declared an international public health emergency, the science related to ZVI was intensified in endemic and non-endemic countries. The evolution of the study field over a short period evolved from epidemiology and entomology to cover clinical and diagnostic aspects of the virus. Subsequently, laboratory-based studies took over, with an emphasis on molecular and cellular biology. This process has mobilized scientific efforts and resources from different countries, institutions and research teams from different scientific areas. In a very short time, complex interactions among these teams from different scientific areas have taken place. An analysis of articles published on this subject in this period provides important insights into our understanding of the path of the scientific response to ZVI.

The strong collaboration between the USA and countries from different continents (e.g., Brazil, Canada, Germany and Australia), showed that the USA is the main contributor in terms of collaboration and suggests its key research capacity in the field. Among the possible reasons why the USA is found to be such a prominent publication country as well as a central player in terms of international collaboration in Zika, might be the fact that it is the top R&D funder for neglected diseases [20]. Furthermore, this country has considerable knowledge accumulation in tropical diseases, including the Department of Defence, which has been one of the main supporters of scientific research in this area [21,22].

By using the International Collaboration Index (ICI) and the International Collaboration Factor (ICF), we were able to examine the influence of the countries by taking into consideration the number of countries they have contributed with and their ratio of international collaboration over time. The most collaborative countries in relative terms are in Europe. The high levels of the international collaboration of England may be explained by the significant recent investment made available by the British government and philanthropic funding agencies for the research community aiming to respond to the challenges posed by ZVI [23]. The same can be said for the European Union, which counts on four large research consortia (ZikaPlan, ZikAction, ZikAlliance and ZikaVax) coordinated by European organizations in association with research institutions from different regions across the world [24].

Also, the potential threat of a Zika epidemic could partially explain the increased country collaboration over time. When the first case of Zika virus infection was confirmed in the Northeast region of Brazil, in May 2015, an epidemiological alert by the Pan American Health Organization (PAHO)/World Health Organization (WHO) was released. The alert recommended Member States to establish and maintain a capacity for ZVI detection, clinical management and an effective public communication strategy [25]. In November 2015, Brazil declared ZVI a national public health emergency and three months later, the World Health Organization declared it a PHEIC [26]. We suggest that a causal effect between the alert/emergency and the increased number of collaborations took place during this period considering the knowledge gaps in understanding ZVI.

Brazil is a country with a vast scientific literature on dengue virus transmission, among other arboviruses. However, the results indicate that it had carried out almost no research on Zika virus before the start of the epidemic. Most of the entomological studies had taken place in countries such as Italy, India and Australia. The scarce research resources for research and the virtual inexistence of Zika virus in the Western Hemisphere until 2014 could justify this finding.

One of the first steps for health surveillance is to take the kind of disease into consideration. In this sense, research into the clinical aspects of Zika helps in the characterization of the disease. At the end of 2015, Zika infection and especially microcephaly appeared as a new problem to be tackled. Questions regarding the clinical aspects and the epidemiology were the main focus at this moment. Physicians might argue: “What is the Diagnosis?”, “What are the physical pathological mechanisms of the virus/disease?”, “What is the case definition?”, putting the focus on physiopathology.

These questions boosted basic science, since research on the clinical aspects and epidemiology depends basically on people and methods, while applied science builds upon cellular and molecular biology, depending on expensive equipment and other technologies and qualifications. This scenario gives an academic advantage to countries with a higher technological density. We saw Cluster 1 “Clinical Aspects” in 2016 being superseded by Clusters 5 and 6 (“Cellular Biology” and “Molecular Biology”) in the following years. It also gives clues to understanding the dominant participation of countries such as the USA and China in the clusters related to cellular and molecular biology.

Nevertheless, these results have some limitations as our source of information was the Web of Science, which could have excluded articles not indexed in this database. All in all, this study, with its strengths and limitations, shows that science has values, interests and practical applications. The fulfilling of the research objectives are thus strategic in the sense that it could aid the dissemination of research, appropriation and the use of the knowledge produced within the framework of scientific research to support strategies for the control and prevention of the ZVI.

However, in the Global Health arena, it is known that inequities in science and technology among high and low and middle-income countries area default in the historical trajectory. Since the Zika outbreak, Brazil has played an important and essential role in developing research and publishing about ZVI and its repercussions. However, regarding ZVI scientific production, we can ask how sustainable the Brazilian prominence in time is. Until when will Brazil be an almost mandatory partner to develop research and publishing on this matter? How to improve more equitable and sustainable scientific production in the so-called “Global South”? Challenging questions yet to be considered in further studies.

## Conclusions

In this study, we have demonstrated that the scientific global research path in response to the public health emergency of ZVI can be traced with data from indexed articles. It shows the value of the study of scientific networks to help to understand science production in response to a PHEIC. As soon as the public health emergency of ZVI started, our findings showed a first phase of the scientific inquiry focused on entomological and epidemiological studies. It was followed by a second phase with a dominance of studies on the diagnosis and clinical aspects of the ZVI, and by a third and fourth phase focused on molecular and cellular biology. We also found that Brazil has played an important role not only in the rapid response to the epidemic but also in the capacity to produce research and to evoke international collaboration within a short period with high-income countries such as the USA and England.

## Data Availability

All data are fully available without restriction.
All relevant data are within the manuscript and its Supporting Information files.

## Acknowledements

We would like to thank all the specialists that evaluate the clusters established in our study.

This work is supported by the Center of Data and Knowledge Integration for Health (CIDACS) through the Zika Platform - a long-term surveillance platform for the Zika virus and microcephaly, Unified Health System (SUS) - Brazilian Ministry of Health.

This work was partially supported by the European Union’s Horizon 2020 Research and Innovation Programme under ZIKAlliance Grant Agreement no. 734548, and by the Oswaldo Cruz Foundation/ Vice-Presidency of Research and Biological Collections-Fiocruz/ VPPCB and the Newton Fund/ British Council.

